# The National Psychiatric Morbidity Survey of Pakistan (2022): Prevalence, socio-demographic and disability correlates

**DOI:** 10.1101/2024.07.08.24310056

**Authors:** R Rahman, D Sheehan, A Javed, SH Ahmad, K Shafiq, U Kanwal, MI Afridi, AT Nizami, G Rasool, R Taj, M Ansari, M Naim, S Farooq, A Memon, MS Karim, Y Hana, M. Ayub

## Abstract

**Background:** National psychiatric morbidity surveys have shown a wide range of prevalence of psychiatric disorders across different countries. Pakistan with its sociocultural and ethnic diversity, has the fifth largest population in the world. There was no prior high-quality nationally representative data on the prevalence of psychiatric disorders and their socio-demographic correlates for Pakistan. To fill this gap in the planning of mental health services, the Pakistan Psychiatric Society conducted the National Psychiatric Morbidity Survey (NPMS) of Pakistan, in the years 2019-2022.

**Aim:** To estimate the prevalence and socio-demographic correlates of psychiatric morbidity in a representative sample of Pakistan.

**Methods:** The cross-sectional NPMS collected data from the four provinces of Pakistan. After selection through a three-stage, stratified, random cluster sampling technique we interviewed 17,773 adults above the age of 18. We used the MINI International Neuropsychiatric Interview (MINI Version 7.0.2) to evaluate psychiatric morbidity. Current and lifetime precise and weighted prevalence is reported according to ICD-10 (International Classification of Disease-10^th^ version). We used multivariate logistic regression to investigate the association between the risk of psychiatric illness and sociodemographic variables. National Bio-ethic Committee of Pakistan granted approval of survey.

**Results:** The lifetime and current weighted prevalence of all psychiatric disorder is 37.91% (95% Confidence Interval (CI) =37.22-38.59) and 32.28% (95% CI=31.62-32.94) respectively. The weighted prevalence of common psychiatric disorders in Pakistan included Mood Disorders (F30-F39; 19.62%), Neurotic and Stress-related Disorders (F40 F48; 24.81%), Psychotic Disorders (F20-F29; 4.52%) and Mental and Behavioural Problems due to Psychoactive Substance use (F10-F19; 0.85%). The psychiatric disorders had an association with age, female gender, urban living, lower income and being divorced. Among participants, 6.17% acknowledged suicidality in the past month, while 1.05% acknowledged a lifetime suicide attempt.

**Conclusion:** The NPMS is the first nationally representative study of psychiatric morbidities in Pakistan. The data from this survey can be utilized for designing and implementing mental health services and support programmes in the country.

## Introduction

Pakistan is a resource limited, lower-middle-income country (LMIC) where like other low and middle-income countries psychiatric disorders are highly prevalent^1^. According to World Health Organization (WHO), 24 million people in Pakistan need psychiatric help^2^. The lack of resources to meet this need is exemplified by 0.19 psychiatrists per 100,000 population, the lowest in the world^2^. Psychiatric disorders are responsible for more than 4% of the total disease burden^2^. The appreciation of the impact of psychiatric disorders is important as they are linked to substantial disability, reduced productivity, low quality of life and worsening socioeconomic circumstances^3^. Our literature search did not reveal any study of the prevalence of psychiatric disorders in a nationally representative sample in Pakistan. Small-scale local surveys have shown widely varying estimates of psychiatric morbidity ^5,6,7,8,9,10,11^. A systematic review^12^ reported a prevalence rate for psychiatric morbidity ranging from 10% to 66%^13^. These figures are higher as compared to other developing countries with similar socioeconomic indicators^6,7,8,14,15^. The significant difference in prevalence among different studies is due to variation in the size of sample, the catchment area of study, operational definitions of psychiatric morbidity, study tool, methodology of study, response rates and quality of monitoring^16.17^. Another estimate of psychiatric disorders is based on WHO-EMRO region country profile statistics that show that 10–16% of Pakistan’s general population has mild to moderate psychiatric illnesses, and 1% has severe mental illnesses^18,19^. In the presence of varied nationwide prevalence estimate of the different psychiatric disorders it will be difficult in Pakistan to implement an affordable, egalitarian, integrated, and data-driven mental healthcare strategy.

There is growing evidence of a link between mental health conditions and non-communicable diseases (NCDs)^20^. The agenda-2030 of United Nation Organization (UNO) to achieve sustainable Developmental Goals (SDG) also focuses on NCD and has included mental health (SDG # 3) and mental disabilities (SDG # 4,8,10 & 11)^21^. To achieve this agenda of UNO, mental health is being integrated into the future health policies of Pakistan (National Health Vision)^22^. Many legislators, experts, and policymakers believe that burden, distribution, and pattern data at the national level is necessary for developing and implementing evidence-based mental healthcare programmes.

Considering the aforementioned facts, the Pakistan Psychiatric Society (PPS), an representative elected body of Pakistani psychiatrists, planned to conduct the National Psychiatric Morbidity Survey (NPMS) in a representative sample from Pakistani population, to investigate the prevalence rates of psychiatric disorders and their socioeconomic correlations and disability levels^23^. We used a sound methodology that addresses some of the shortcomings of the previous work. The data of this survey would help in planning and improving mental health in Pakistan. The findings of NPMS are being described in this article, with a focus on the following objectives:

- To assess the prevalence of common psychiatric disorders in a representative sample in Pakistan.
- To assess the relationship between psychiatric morbidity and different demographic variables (gender, age, education, employment, residence and income).
- To evaluate suicidal ideation and behaviours.

## Methodology

A concise synopsis of the NPMS survey methodology can be found at (https://pakistanpsychiatricsurvey.org/)^24^ and a summary is given here. The survey was overseen by an Executive Committee of Survey which included national and international experts. The Committee had diverse expertise and included, professionals, members of PPS, epidemiologists, subject experts in mental health, demographers, and statisticians. The approval of NPMS was given by the National Bioethics Committee of Pakistan (letter No. 4-87/NBC-268/171/370, dated November 11, 2017) as well as the institutional review board of the Dow University of Health Sciences. To maintain uniformity in the datagathering process, the Executive Committee developed the Master Protocol and Operational instructions for data collection and fieldwork. A pilot study was carried out in a suburb of Karachi to standardize the process of methodology and identify any difficulty in fieldwork. The main issue identified during the pilot study was the non-acceptance of male interviewers by some communities. As a result, we decided to send one male and one female interviewer to each household to conduct the interviews. Informed consent was obtained from the head of the household and each participant.

The sample was randomly identified by Pakistan Bureau of Statistics (PBS), from all over the country, to represent various geographical regions and to ensure cultural diversity. PBS is Pakistan’s prime state agency, responsible for collecting, compiling and disseminating reliable statistical information timely to policy makers, planners and researchers^25^. A three-stage, stratified, random cluster sampling procedure was used for the survey. In the first stage, PBS randomly selected enumeration blocks based on probability proportionate to size. According to the Census of Pakistan 2017^25^, the urban population was divided into enumeration blocks each having 200 to 300 households, while in the rural areas each village is an independent enumeration block. This selection of enumeration blocks helped in generating data at the provincial and national levels. The sample frame for the ensuing systematic random sampling of households was formed by serially listing and numbering every household inside the selected block. Using the Kish method^26,27^, primary respondents above 18year from the households were selected for interview, who comprehend the basic rational of research and give informed consent. Where no participant fulfilling the inclusion criteria was available in the selected house then the next house in the serial list was approached. The sociodemographic data of all individuals living together for at least six months and sharing the same kitchen were collected. Guests visiting temporarily were excluded.

Using standard statistical techniques for estimating sample size in population-based surveys, the PBS calculated the sample size of 11180 adults (>18 years of age), spread across all four provinces {based on prevalence 34%, design effect 4 (d=margin of error 4%), level of confidence 95% non-response 3%}. To increase the sample size because of the convenience of getting a larger sample size in these regions, the sample was doubled in Sindh and Punjab. In order to identify psychiatric disorders in the study population, the Mini International Neuropsychiatric Interview (MINI) schedule Version 7.0.2^28^ was used. The MINI is a structured diagnostic interview tool used to assess, document and confirm the presence of common psychiatric disorders in community-based epidemiological studies. The MINI is compatible with both DSM-5 (Diagnostic and Statistical Manual of Mental Disorders, fifth edition) and the ICD-10 (International Classification of Diseases, tenth revision)^29,30,31^. The MINI has built-in diagnostic algorithms that produce diagnoses that are in alignment with the DSM-5 and ICD-10. When compared to other instruments, the MINI was compact, simple to use, efficient in terms of administration time, valid and reliable, and helpful in both clinical and community contexts. The MINI was translated into Urdu, the national and most widely used language in Pakistan. It is spoken in all provinces of Pakistan. Although suicidality is not included as a diagnostic entity in different classifications of psychiatric disorders, but this predictor of psychiatric mortality is being assessed through the module-B of MINI. We used the the module-B of MINI to assess the different aspects of suicidal behaviours in detail that is specifically helpful in predicting suicidal attempt.

The Sheehan Disability Scale^32^ was used to assess disability due to mental disorders, in three areas including family life and home responsibilities, work and social life. Dow University of Health Sciences translated and piloted all study instruments used in NPMS. For quality assurance the process of back translation was adopted in which conceptual translation of MINI in Urdu language was literally back translated into English by a third-party linguist and any deviation from the original version were discussed and reconciled.

We hired field workers preferably with a background in psychology/sociology/social work or related fields to conduct the in-person interviews. We provided systematic and thorough training in the administration of the study tools and in how to follow the survey methodology to the field workers. Pre & post training tests were used for quality assurance.

In order to ensure the collection of high-quality data, a reliable three-tier monitoring system was adopted that included spot checks, supervised field trips, and weekly and monthly review meetings. Every day, the data gathered from each location were uploaded to a central data pool. The final dataset was created by compiling and cleaning the collected data. The current or point prevalence was calculated for all psychiatric disorder while life-time prevalence was calculated for all-psychiatric morbidities, mood disorders (F30–F39), psychotic disorders (F20–F29), neurotic and stress-related disorders (F40–F48), other anxiety disorders (F41) and panic disorder (F41.0). The all-psychiatric morbidity was defined as a disorder that was identified by the MINI instrument and that fell under the ICD-10 Diagnostic Criteria for Research (DCR) (F10–F19, F20–F29, F30–F39, and F40–F48). Tobacco related psychological conditions are not included in the definition of psychiatric morbidity so their prevalence rates were not calculated. Although suicidality is also not included as an ICD category it is analyzed as it a major psychiatric “condition” and a leading cause of mortality from psychiatric disorders, so its data is important for our country. Because of the unequal probability of selection due to more data being collected from Sindh and Punjab and also due to the nonresponse rate, we computed weighted prevalence estimates of psychiatric morbidities as percentage and confidence intervals (CIs). We used binary and multivariate logistic regression with normalised weights to investigate the relationship between psychiatric disorders as a dependent variable and several socio-demographic characteristics (gender, age, education, marital status, occupation, income, religion and geographic location) as independent variables. The risk of developing a psychiatric disease for each participant in the selected group was compared to the risk in the reference group using adjusted odds ratios that were generated using the model. All analyses were performed using SPSS (version-26)^33^.

## Results

In total 20833 households were approached as per study procedure in the four provinces of Pakistan, and 17,773 eligible participants were interviewed (response rate, 86.2%). The reasons for non-response were a locked house (n=628), non-residential building (n=144), no adult above 18 years was available at home (n=369), local area or residential security staff did not allow the team to work (n=13), refusal at door (n=1329), house doors were not opened (n=339), no one fulfilling the inclusion criteria was present at home (n=54). Less than 0.9% (n=184) participants refused to continue the interview.

Sociodemographic characteristics of sample are given in Table-1 that shows that one-third of the study sample (33.6%) consisted of respondents who were between the ages of 18 and 29. Females constituted 50.8% of the sample, while 63.8% of the study participants were from rural areas. While, 33.7% of the study subjects were classified as “not literate,” and 81.1% were married.

**Table 1.** Sociodemographic characteristics of study population (N=17773).

| Characteristics |  | n | Frequency (%) |
| --- | --- | --- | --- |
| Province | Punjab | 7680 | (43.2) |
|  | Sindh | 5680 | (32.0) |
|  | KPK | 2524 | (14.2) |
|  | Baluchistan | 1889 | (10.6) |
| Gender | Female | 9023 | (50.8) |
|  | Male | 8750 | (49.2) |
| Age (Years) | 18-29 | 5970 | (33.6) |
|  | 30-39 | 5500 | (30.9) |
|  | 40-49 | 3590 | (20.2) |
|  | 50-59 | 1676 | (9.4) |
|  | 60+ | 1037 | (5.8) |
| District | Rural | 11338 | (63.8) |
|  | Urban | 6435 | (36.2) |
| Marital Status | Married | 14419 | (81.1) |
|  | Unmarried | 2844 | (16.0) |
|  | Widowed | 390 | (2.2) |
|  | Separated | 73 | (0.4) |
|  | Divorced | 47 | (0.3) |
| Religion | Islam | 17314 | (97.4) |
|  | Hindu | 255 | (1.4) |
|  | Christianity | 204 | (1.1) |
| Level of Education | No schooling | 5991 | (33.7) |
|  | Primary/Middle (5 or 8 years) | 4496 | (25.3) |
|  | Matric (10 years) | 3347 | (18.8) |
|  | Intermediate (12 years) | 1901 | (10.7) |
|  | Graduate & above (14 years & above) | 1552 | (8.0) |
|  | Others | 486 | (2.7) |
| Occupation | Working (on job) | 7046 | (39.6) |
|  | Jobless | 4236 | (23.8) |
|  | Homemaker | 4070 | (22.9) |
|  | Students | 379 | (2.1) |
|  | Retired | 143 | (0.8) |
|  | Other | 1899 | (10.7) |
| Family Type | Nuclear | 11800 | (66.4) |
|  | Joint - more than one family related families living together | 5973 | (33.6) |
| Income per month (rupees) | 5000 or less (USD 30 or less) | 1678 | (9.4) |
|  | 5001-8000 (USD 30-55) | 1218 | (6.9) |
|  | 8001 -15000 (USD 30-35) | 3766 | (21.2) |
|  | 15001-25000 (USD 55-90) | 4384 | (24.7) |
|  | 25001-45000 USD 90-160) | 3026 | (17.0) |
|  | 45001 or more | 1273 | (7.2) |
|  | No answer | 2428 | (13.7) |

Male and female mean ages were 36.25±12.70 and 35.74±11.98 years, respectively. The research population’s age and gender distribution roughly matched the national values (Census-2017)^34^, with the exception of a slightly greater percentage of older people (>60 years old; 5.8% in the sample compared to 5.22% in the Census 2017). The NPMS research population’s age, gender, marital status, and literacy level distributions closely matched the proportions from the 2017 Census. Groups based on the greatest self-reported monthly income shows that monthly income of PKR 15,001 to 25,000 (USD 55 to 90) was 24.7% of the sample, followed by 8,001-15,000 (USD 30 to 55) that was 21.2% & 25,001 to 45,000 (USD 90 to 160) per month that was 17.0% (Table-1).

The weighted prevalence of various psychiatric morbidities is shown in Table-2. “All mental morbidities” was projected to have a weighted lifetime prevalence of 37.91% (95% CI=37.22, 38.59) and the current prevalence of 32.28% (95% CI= 31.62, 32.94). The prevalence of mood disorders (F30–F39) was 11.57% current and 19.62% lifetime, with depressive disorders (F32–F33) being the most common subgroup. Of depressive disorders, the current prevalence (F32–F33) was over half of the lifetime prevalence. The projected lifetime prevalence of stress-related and neurotic disorders (F40–F48) was 24.81%. (Table 2).

**Table 2.**
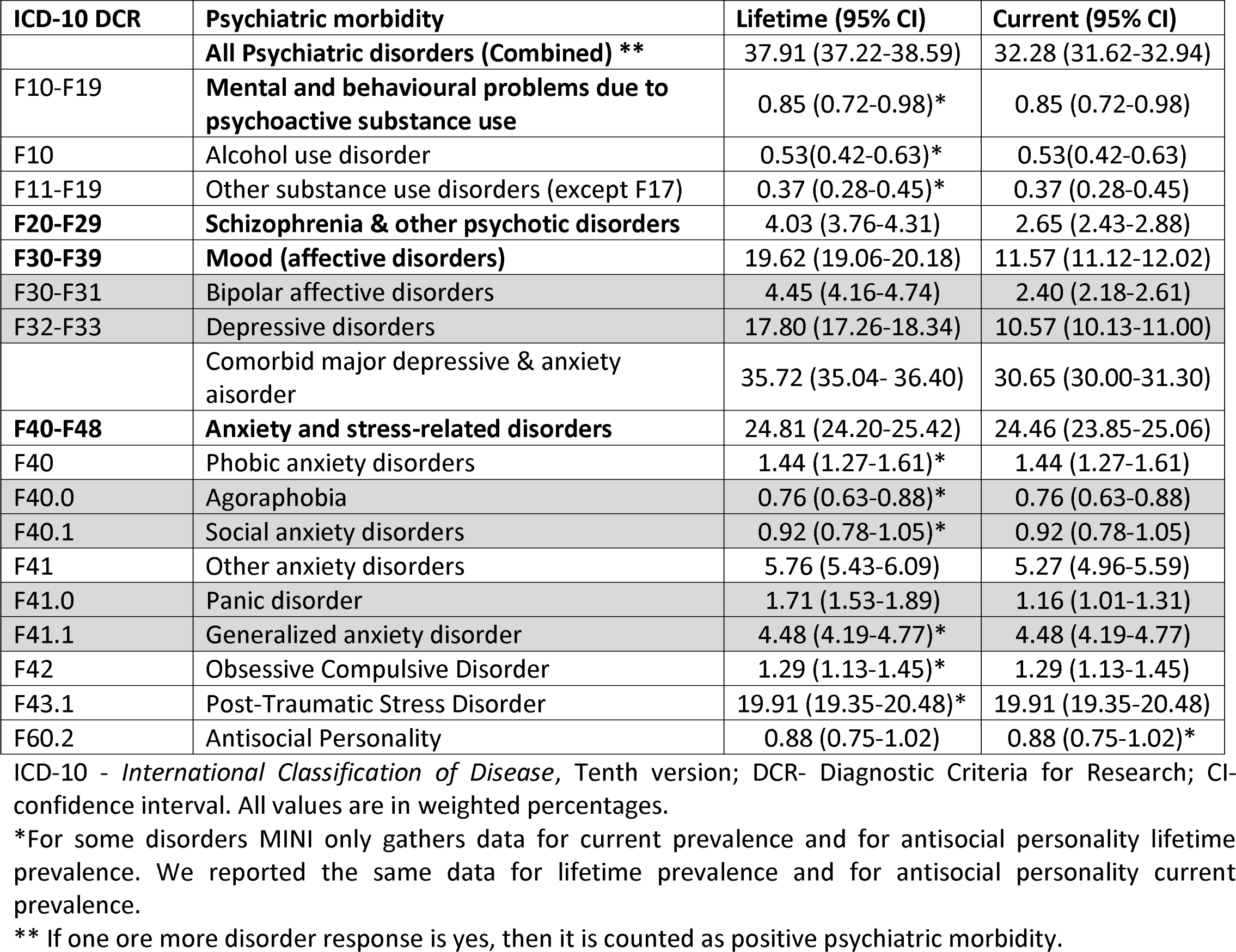
Prevalence of psychiatric morbidity as per ICD-10 DCR among adults (18+ years) (*n* = 17,773).

| ICD-10 DCR | Psychiatric morbidity | Lifetime (95% CI) | Current (95% CI) |
| --- | --- | --- | --- |
|  | **All Psychiatric disorders (Combined) \*\*** | 37.91 (37.22-38.59) | 32.28 (31.62-32.94) |
| F10-F19 | **Mental and behavioural problems due to psychoactive substance use** | 0.85 (0.72-0.98)* | 0.85 (0.72-0.98) |
| F10 | Alcohol use disorder | 0.53(0.42-0.63)* | 0.53(0.42-0.63) |
| F11-F19 | Other substance use disorders (except F17) | 0.37 (0.28-0.45)* | 0.37 (0.28-0.45) |
| **F20-F29** | **Schizophrenia & other psychotic disorders** | 4.03 (3.76-4.31) | 2.65 (2.43-2.88) |
| **F30-F39** | **Mood (affective disorders)** | 19.62 (19.06-20.18) | 11.57 (11.12-12.02) |
| F30-F31 | Bipolar affective disorders | 4.45 (4.16-4.74) | 2.40 (2.18-2.61) |
| F32-F33 | Depressive disorders | 17.80 (17.26-18.34) | 10.57 (10.13-11.00) |
|  | Comorbid major depressive & anxiety disorder | 35.72 (35.04- 36.40) | 30.65 (30.00-31.30) |
| **F40-F48** | **Anxiety and stress-related disorders** | 24.81 (24.20-25.42) | 24.46 (23.85-25.06) |
| F40 | Phobic anxiety disorders | 1.44 (1.27-1.61)* | 1.44 (1.27-1.61) |
| F40.0 | Agoraphobia | 0.76 (0.63-0.88)* | 0.76 (0.63-0.88) |
| F40.1 | Social anxiety disorders | 0.92 (0.78-1.05)* | 0.92 (0.78-1.05) |
| F41 | Other anxiety disorders | 5.76 (5.43-6.09) | 5.27 (4.96-5.59) |
| F41.0 | Panic disorder | 1.71 (1.53-1.89) | 1.16 (1.01-1.31) |
| F41.1 | Generalized anxiety disorder | 4.48 (4.19-4.77)* | 4.48 (4.19-4.77) |
| F42 | Obsessive Compulsive Disorder | 1.29 (1.13-1.45)* | 1.29 (1.13-1.45) |
| F43.1 | Post-Traumatic Stress Disorder | 19.91 (19.35-20.48)* | 19.91 (19.35-20.48) |
| F60.2 | Antisocial Personality | 0.88 (0.75-1.02) | 0.88 (0.75-1.02)* |

ICD-10 - *International Classification of Disease*, Tenth version; DCR- Diagnostic Criteria for Research; CI- confidence interval. All values are in weighted percentages.
\*For some disorders MINI only gathers data for current prevalence and for antisocial personality lifetime prevalence. We reported the same data for lifetime prevalence and for antisocial personality current prevalence.
\*\* If one ore more disorder response is yes, then it is counted as positive psychiatric morbidity.

Currently suicidality is not a diagnostic entity, but its identification is important for prevention strategies, so the module-B of MINI assesses the different aspects of suicidal behavior ranging from non-suicidal deliberate self-harm to active or passive suicidal ideation, suicidal intent, plan and attempt. 6.17% acknowledged some type of suicidality in the past month. 1.05% acknowledged a lifetime suicide attempt. The details of the full spectrum of suicidal phenomena are summarized in Table-3.

**Table 3.** Prevalence of suicidal phenomenon among adults (18+ years) (*N* = 17,773).

| Suicidal phenomenon in past one month |  | Frequency | % (Confidence Interval) |
| --- | --- | --- | --- |
| Suicidality: At least one response in B-module is yes except B1 |  | 1301 | 7.32 (6.94 – 7.70) |
| Suicidal Accidents involving suicidal intent: – B1a |  | 177 | 1.00 (0.85 – 1.14) |
| Passive Suicidal Ideation: – B2 |  | 735 | 4.14 (3.84 – 4.43) |
| Active Suicidal Ideation: – B3 |  | 570 | 3.21 (2.95 – 3.47) |
| Psychotic Suicidality: – B4 – a voice telling you to kill yourself |  | 109 | 0.61 (0.50 – 0.73) |
| Suicidal Dreams: – B4 – a dream with suicidal content |  | 88 | 0.50 (0.39 – 0.60) |
| Impulsive Suicidality: – B12 |  | 179 | 1.01 (0.86 – 1.15) |
| Suicidal Plan: – B5 and/or B6 and/or B7 and/or B8 and/or B9 |  | 325 | 1.83 (1.63 – 2.03) |
| Suicidal Intent: – B10 or B11 |  | 365 | 2.05 (1.85 – 2.26) |
| Suicidal Preparatory Behavior: – B9 or B14 |  | 232 | 1.31 (1.14 – 1.47) |
| Suicide Attempt: – B16 |  | 186 | 1.05 (0.90 – 1.20) |
| Suicide attempt lifetime: – B18 |  | 186 | 1.05 (0.90 – 1.20) |
| Suicidality in the past month: -<br>B1b/B2/B3/B4/B5/B6/B7/B8/B9/B10/B11/B12/B14/B16. |  | 1097 | 6.17 (5.82 – 6.53) |
| Death by Suicide: N/A |  |  |  |
| Non-suicidal self-injury: – B15 |  | 151 | 0.85 (0.71 – 0.98) |
| Usual time spent per day with<br>any suicidal impulses,<br>thoughts or actions: – B17a | Up to 120 minutes | 448 | 2.52 (2.29 – 2.76) |
|  | 120 to 360 minutes | 379 | 2.13 (1.93 – 2.36) |
|  | More than 360 minutes | 16 | 0.90 (0.05 – 0.15) |

The age group of 50–59 years old had the highest lifetime (63.68%) and current prevalence (60.67%) of psychiatric disorders. Among (60 years of age and over), lifetime and current prevalences were 59.19% and 56.73% respectively.

Females had a greater lifetime (38.50% vs 37.31) and current (33.46% vs 31.07%) prevalence of “all mental morbidities.” In comparison to rural areas (37.72% and 31.51%), the prevalence of psychiatric morbidity (both lifetime and current) was greater in urban areas (39.31% and 33.84%). Divorced, widow and separated people had a greater prevalence of mental illness (lifetime = 71.35%, 66.36% & 51.85% current = 63.81%, 60.86% and 40.09%) than married and unmarried (lifetime = 39.36% & 26.15% current = 34.14% and 18.64%). Households in the lower income quintile had higher rates of mental illness. People who spent more years in school had a lower prevalence of mental illness. For example, in those with no formal schooling the prevalence was 42.84%, in those with five years of schooling it was 40.88%. It decreased to 25.47% among the participants who had twelve years of schooling. (Table 4).

**Table 4.** Socio-demographic features and Odds for psychiatric morbidity (N=17,773).

| Characteristic | Weighted Prevalence |  | Risk for current mental morbidity |  |  |
| --- | --- | --- | --- | --- | --- |
|  | Lifetime (95% CI) | Current (95% CI) | Crude OR | Adj. OR | 95% CI |
| Psychiatric morbidity | 37.91 (37.22-38.59) | 32.28 (31.62-32.94) |  |  |  |
| Age group (years) |  |  |  |  |  |
| 18–29 | 23.25 (22.24-24.26) | 15.77 (14.90-16.64) | Ref | Ref |  |
| 30–39 | 29.33 (28.18-30.48) | 22.67 (21.61-23.73) | 1.57** | 1.55** | 1.41 - 1.71 |
| 40–49 | 59.83 (58.26-61.40) | 57.04 (55.45-58.63) | 7.09** | 7.26** | 6.56 - 8.05 |
| 50–59 | 63.68 (61.42-65.93) | 60.67 (58.38-62.96) | 8.24** | 8.32** | 7.32 - 9.45 |
| ≥60 | 59.19 (56.24-62.14) | 56.73 (53.75-59.70) | 7.00** | 6.54** | 5.62 - 7.61 |
| Sex |  |  |  |  |  |
| Male | 37.31 (36.33-38.28) | 31.07 (30.14-32.00) | Ref | Ref | Ref |
| Female | 38.50 (37.53-39.46) | 33.46 (32.52-34.39) | 1.11** | 0.75** | 0.69 - 0.82 |
| Residence |  |  |  |  |  |
| Urban | 39.31 (38.11-40.51) | 33.84 (32.68-35.01) | Ref | Ref | Ref |
| Rural | 37.22 (36.39-38.06) | 31.51 (30.71-32.31) | 0.90* | 0.83** | 0.77 - 0.89 |
| Education |  |  |  |  |  |
| Uneducated | 42.84 (41.63-44.05) | 39.06 (37.87-40.24) | Ref | Ref | Ref |
| Primary/Middle (5 or 8 years) | 40.88 (39.49-42.28) | 35.13 (33.78-36.49) | 0.84** | 1.09 | 0.99 - 1.19 |
| Matric (10 years) | 33.57 (32.03-35.10) | 28.05 (26.59-29.51) | 0.61** | 0.93 | 0.84 - 1.03 |
| Intermediate (12 years) | 25.47 (23.61-27.32) | 18.87 (17.20-20.53) | 0.36** | 0.65** | 0.57 - 0.75 |
| Graduate or Above (14 years & above) | 32.84 (30.55-35.13) | 24.61 (22.52-26.71) | 0.51** | 0.88 | 0.79 - 1.02 |
| Others | 44.75 (40.87-48.63) | 30.43 (26.84-34.02) | 0.68** | 0.89 | 0.73 - 1.08 |
| Occupation |  |  |  |  |  |
| Working (on job) | 32.79 (31.71-33.87) | 27.38 (26.36-28.41) | Ref | Ref | Ref |
| Jobless | 36.44 (35.02-37.86) | 31.09 (29.72-32.46) | 1.20** | 1.35** | 1.23 - 1.49 |
| Homemaker | 43.75 (42.32-45.18) | 38.98 (37.58-40.39) | 1.69** | 2.52** | 2.26 - 2.81 |
| Students | 33.34 (28.83-37.84) | 23.86 (19.79-27.93) | 0.83 | 2.36** | 1.84 - 3.04 |
| Retired | 66.15 (58.44-73.87) | 62.66 (54.78-70.55) | 4.45** | 2.91** | 2.00 - 4.22 |
| Others | 43.83 (41.84-45.81) | 35.91 (33.99-37.82) | 1.49** | 1.88** | 1.68 - 2.11 |
| Marital status |  |  |  |  |  |
| Unmarried | 26.15 (24.59-27.70) | 18.64 (17.26-20.21) | Ref | Ref | Ref |
| Married | 39.36 (38.59-40.12) | 34.14 (33.40-34.88) | 2.26** | 1.01 | 0.90 - 1.13 |
| Separated | 51.85 (42.09-61.60) | 40.09 (30.52-49.66) | 2.92** | 1.56 | 0.98 - 2.46 |
| Divorced | 71.35 (57.88-84.81) | 63.81 (49.50-78.12) | 7.70** | 4.34** | 2.23 - 8.43 |
| Widowed | 66.36 (61.52-71.20) | 60.86 (55.86-65.86) | 6.79** | 1.43* | 1.11 - 1.85 |
| Income quintile |  |  |  |  |  |
| 5000 or less (USD 30 or less) | 42.90 (40.57-45.22) | 37.83 (35.55-40.10) | Ref | Ref | Ref |
| 5001-8000 (USD 30-55) | 42.69 (39.89-45.48) | 36.26 (33.55-38.98) | 0.93 | 0.97 | 0.82 - 1.15 |
| 8001 -15000 (USD 30-35) | 37.47 (35.94-39.00) | 33.90 (32.40-35.40) | 0.84* | 0.80* | 0.70 - 0.92 |
| 15001-25000 (USD 55-90) | 38.15 (36.76-39.54) | 32.32 (30.98-33.66) | 0.78** | 0.81* | 0.71 - 0.92 |
| 25001-45000 USD 90-160) | 35.78 (34.17-37.39) | 29.30 (27.77-30.83) | 0.68** | 0.72** | 0.63 - 0.83 |
| 45001 or more | 32.50 (29.93-35.07) | 26.60 (24.18-29.02) | 0.59** | 0.65** | 0.54 - 0.78 |
| No Answer | 38.00 (36.29-39.70) | 31.16 (29.53-32.79) | 0.74** | 0.68** | 0.59 - 0.78 |

\*p<0.05. \*\*p< 0.01. CI: confidence interval; Crude OR: crude odds ratio, Adj.: adjusted.

The Odds of psychiatric illness were 7 to 8 times higher in old age above 40 years compared to young individuals between the ages of 18 and 29 (Table 4). Odds for psychiatric morbidity was nearly three times higher in retired persons (Adj. OR=2.9, 95% CI=2.00, 4.22; *p* < .001). People who were divorced were at a higher Odds of psychiatric disorders (Adj. OR = 4.34, 95% CI = 2.23, 8.43; *p* < .001). Odds for psychiatric morbidity was lower among individuals with highest income as compared to lower income persons.

Female (Adj. OR= 0.75, 95% CI = 0.69, 0.82; p <.001) had lesser risk of psychiatric morbidity compared to males, and Rural inhabitants (Adj. OR = 0.83, 95% CI = 0.77, 0.89; p <.001) had lesser risk compared to the urban population. More time in education was associated with less risk of reporting psychiatric illness. The odds of psychiatric illness were higher for those in the lowest quintile of income than for those in the top quintile. The findings confirm that male gender, less education, low income, living in an urban area, and being divorced or separated are all linked to psychiatric morbidity.

In this survey functional impairment due to psychiatric disorders, in different areas of life including; family life, home responsibilities, social and occupational functioning, was assessed through the Sheehan Disability Scale^32^, the association of functional disability with psychiatric disorders is shown in the Table-5:

**Table 5:** Level of disability in patients with psychiatric morbidity.

| ICD-10 DCR | Mental morbidity | Life time SDS<br>Mean ± SD | Current SDS<br>Mean ± SD |
| --- | --- | --- | --- |
| | **All mental morbidity (Combined)** | 18.99 ± 13.14 | 19.22 ± 12.99 |
| F10-F19 | **Mental and behavioural problems due to psychoactive substance use** | 20.16 ± 11.40 | |
| F10 | Alcohol use disorder | 23.71 ± 11.37 | |
| F11-F19 | Other substance use disorders (except F17) | 16.64 ± 10.27 | |
| **F20-F29** | **Schizophrenia & other psychotic disorders** | 22.64 ± 11.61 | 23.29 ± 11.66 |
| **F30-F39** | **Mood (affective disorders)** | 19.23 ± 12.84 | 18.79 ± 12.28 |
| F30-F31 | Bipolar affective disorders | 23.75 ± 11.12 | 23.54 ± 10.71 |
| F32-F33 | Depressive disorders | 18.79 ± 12.89 | 18.30 ± 12.24 |
| | Comorbid Major Depressive & Anxiety Disorder | 19.06 ± 13.19 | 19.28 ± 13.02 |
| F40-F48 | **Anxiety and stress-related disorders** | 20.09 ± 13.01 | 20.18 ± 13.0 |
| F40 | Phobic anxiety disorders | 15.50 ± 10.07 | |
| F40.0 | Agoraphobia | 16.99 ± 10.92 | |
| F40.1 | Social anxiety disorders | 14.87 ± 9.02 | |
| F41 | Other anxiety disorders | 20.31 ± 11.87 | 20.63 ± 11.75 |
| F41.0 | Panic disorder | 18.18 ± 12.41 | 15.54 ± 14.39 |
| F41.1 | Generalized anxiety disorder | 21.53 ± 11.32 | |
| F42 | Obsessive Compulsive Disorder | 17.23 ± 10.56 | |
| F43.1 | Post-Traumatic Stress Disorder | 20.88 ± 13.18 | |
| F60.2 | Antisocial Personality | 20.67 ± 10.11 | |

## Discussion

The NPMS is the first national population-based representative survey of psychiatric morbidity in Pakistan (2022).). Its main strength is its ability to address the methodological shortcomings of previous mental health surveys^12^ by using standardised survey protocols together with quality monitoring of data collection. Data were gathered by trained and competent interviewers using a standardized MINI interview schedule. A tiered data monitoring system was implied for quality control. The rigorous methodology and implementation process were adopted.

Because research subjects were interviewed through systematic planned visits and revisits, a high response rate of 86% was achieved in the NPMS. The World Mental Health Survey have response rates ranging from 50.4% (Poland) to 97.2% (Medellín, Columbia), with a pooled response rate of 68.3% across surveys^35^ and it was 79.9% in Nigeria^36^, 88.2% in Germany^37^, 85% in Malaysia^38^, 86.2% in Iran^39^ and 70% in Lebanon^40^. The NPMS offered greater representativeness (external validity) because of the large sample size (17,773 participants from 4 provinces) randomly identified and selected by a robust methodology guided by PBS, which also conducts the census of Pakistan. The sample’s size was determined using a scientific sampling technique (multilevel cluster random sampling with population proportion to size), and it was shown to be representative of the Pakistani population because its age, gender, and rural-urban distribution closely matched Census 2016 proportions.

### Prevalence

The weighted point prevalence of psychiatric morbidity in adults is 31.6%. That means one in three adults in Pakistan is reported to have a mental health Condition at any given time. Our results are lower than those of earlier studies conducted in various regions of Pakistan, where prevalence ranged from 33% to 60%.^12^. The NPMS’s prevalence estimate is greater than that provided in the WHO EMRO region’s country profile data, which show that 10% to 16% of Pakistan’s general population has mild to moderate psychiatric disorders and 1% has severe mental illnesses^18,19^. While point prevalence estimates are important, lifetime prevalence estimates are helpful in planning and providing afflicted individuals with mental health care.

When compared to national-level surveys conducted in other countries the present estimate of the NPMS prevalence is higher than the rates recorded in South Africa (16.5%)^41^, China (17.6%)^42^ and Japan (22%)^43^. In middle- and low-income countries like Malaysia, Nigeria and Iran, the estimates varied from 5.2% to 23.2%. ^37, 38, 39^. The NPMS prevalence rates exceeded the same from England (15.7%).^44^. Our rates exceeded the “12-month prevalence rate of 3.3% and 7.4% mentioned in the WMHS for the African and Asian countries. Nonetheless, NPMS rates for lifetime prevalence of any psychiatric morbidity (37.1%) were within the global range (WMH) of 12-47.4% IRQ 18.1%–36.1%^45^.The possibility of direct comparison of estimates of NMHS with other nation’s data is limited due to differences in culture, values, diagnostic criteria (ICD-10 vs. DSM-IV), age group included in study (18+, 16-65 years) and tool used for study, like the SCID (Structured Clinical Interview for Diagnosis), the CIDI (Composite International Diagnostic Interview), and the MINI (Mini International Neuropsychiatric interview). Despite the differences in socioeconomic environment of the nations, we think NMHS has produced an acceptable and trustworthy estimate. We found variations in prevalence estimates between provinces, which we will report in more depth in our next publication.

The bulk of mental illness was composed of neurotic diseases (F40–F48) and mood disorders (F30–F39). The net weighted prevalence of common psychiatric illnesses (depressive, neurotic and stress-related disorders) was respectively 35.7% & 36.6% for current & lifetime experiences and it constitutes over 95% of all psychiatric morbidities in Pakistan. The majority of severe psychiatric disorders (4.52%) were caused by schizophrenia and other psychotic disorders (4.03%). Although the severe psychiatric disorders including schizophrenia, mania and psychotic depression are less common than other psychiatric disorders, they require more resources due to the nature of these illnesses that have a greater impact on the community.

In this survey the prevalence of depressive disorder is 17.8% that is lower than reported in some previous studies from Pakistan. According to studies & reviews, the prevalence of depression ranged from 22% to 66%^5,13,16^. Methodological differences account for much of this variation. The studies using questionnaires for estimation of prevalence^45,46,47,48^ and studies where participants were screened with a questionnaire and a proportion of high and low scorers was interviewed in the second stage^6,7,8,15^ reported a high prevalence. The likelihood is that these studies were measuring psychological distress as opposed to depressive disorders. This notion is supported by the findings of two studies that used more detailed assessments, which were lower, such as 3.4% across both genders^49,50^. The estimates of this study are within the global range (WMH)) of 12.0%–47.4%^51^

Sheehan has proposed an elaborate Classification Algorithms for Suicidality that provides a granular assessment of suicidal and non-suicidal deliberate self-harming behavior^52^. A clear and relatively comprehensive classification adopted by this approach enhances the reliability and validity of the assessment^53^. The analysis of data according to new classification algorithm revealed that suicidality was present in the form of suicidal ideation (7.3%), plan (1.0%) and attempt (1.0%) that is within the range of suicidal ideation (3.5-11.1%), plan (0.9-9.5%) and attempt (0.3-7.4%) in LMIC (Low- and Middle-Income Countries)^54,55,56^. Further analysis of suicidality in this study during last one month came out to be 6.17% while 1.05% of participant acknowledged a life time suicidal attempt. This information is important as according to WHO data in Pakistan, 8.9 suicides occurred per 100,000 people (male 13.3 % and female 4.3 %) during 2019, and between 15 and 35 people die by suicide every day. This is a rate of one person killing themselves every hour^57^. The amount of time someone spends every day engaging in suicidal impulses, thoughts or actions is an important marker of suicide severity is also analyzed in this study. One third (34.2%) of individuals with suicidal ideation have non-suicidal self-injury. The analysis of suicide behavior in this way will inform the planning of care for individuals at risk of suicide and self-harm.

### Level of disability

In this study we assessed the level of disability or functional impairment in different psychiatric disorders through 12-item SDS (Sheehan Disability Scale) questionnaire^32^. Each item of this scale identifies the level of difficulty, starting with ‘no difficulty’ and including ‘mild’, ‘moderate’, ‘severe’ or ‘extreme’ difficulties. SDS measures functional impairment in three areas including work/school, social life, and home/family responsibilities^58^. It is a simple, brief tool feasible to apply and having good validity & reliability. The higher scores represent severe functional impairments^59^. The results of the SDS demonstrate that all the psychiatric disorders identified in the community sample are associated with moderate to severe levels of functional impairment, not only in work performance, but also in their social life/leisure activities, and their family life and home responsibilities performance. This may be different from hospital population as most of the people are functioning in the community.

### Socio-demographic correlates

There is significant increase in risk of psychiatric morbidity with increasing age especially in people above the age of 40 years. This finding may benefit the service planning where the people with icreasing age may be targeted. It is hard to make any aetiological inference but we can speculate that the people with increasing age has experience the greater social change and they have to provide physical, psychological and financial support to their children and their elders at the same time. The traditional support in the family and community has declined. Additionally, people above the age of 40 years is higher risk of non-communicable diseases including diabetes, hypertension and heart disease. These chronic health conditions have an association with psychiatric disorders.

Although the overall prevalence of psychiatric disorders is slightly high among female (38.5%) as compare to male (37.3%) but adjusted odd ration shows low impact of sex (<1) on psychiatric disorder (Table-4). This finding is similar to other surveys conducted in the general population showing not much difference between males and females in the overall prevalence of mental disorders^59^. But there is difference between men and women in relation to pattern of psychiatric disorders and their symptoms i.e. women suffer from higher rates of depression and anxiety (referred to as internalizing disorders), and men have higher rates of substance abuse and antisocial disorders (referred to as externalizing disorders).^60,61^.

We found that compared to rural areas, the risk of psychiatric morbidity was higher in urban areas (Table 4). This may be due to factors related to the onset of psychiatric illness and better availability of services for early diagnosis in urban areas. Nuclear families, migration, changing lifestyles, competitive life and work environments in cities along with the presence of vulnerable congregations like slums are some of the factors that could be responsible for the higher rate of psychiatric morbidity in urban areas. Factors supporting identification and management of psychiatric illness include; improved services for mental health care, increased awareness of mental health through media and technology^62^. Pakistan is experiencing rapid urbanisation with an increase in the number of people living in cities. In 2017 36.4% (94 million) people were residing in cities and by 2025 50% will be living in cities ^63^. Considering the urbanization trends there is a dire need to strengthen the mental health programmes in cities and to integrate mental health services into other development policies and activities in urban areas.

It is well known that the burden of psychiatric morbidity is higher in low and middle-income nations. The results of the NPMS study confirm that people from economically disadvantaged and socially marginalised groups in society had greater risk and prevalence of psychiatric illness. The socioeconomic environment contributes to the development of common psychiatric disorders^64^. A higher burden of psychiatric morbidity in this population may be caused by a number of factors, including the economic deprivation leading to a heightened vulnerability and restricted access to mental health care that further lowers productivity and family income ^9,65,66^. This heightened socioeconomic burden can be prevented through early intervention in vulnerable groups through partnerships with several government departments, civic society organisations and other stakeholders^64^. Mental health care can be made part of national health initiatives. Increasing rehabilitation and support services available to low-income people and families and implementing initiatives to reduce stigma at every stage would lessen the burden.

Due to growing knowledge about the connection between mental health and NCD (non-communicable diseases), and productivity at work, mental health has become a higher concern in recent years. The establishment of federal and provincial Mental Health Authorities to register and regulate mental health services and handle grievances is a positive step towards the National Health Vision of Pakistan 2016–2025^67^. The evidence-based, valid nationwide data captured by this NPMS can serve as a valuable baseline to evaluate the progress and success of these policies and programs in the future.

### Limitations

Kashmir and Gilgit Baltistan were not included in the NPMS, however, the surveyed provincial population is representative as it covers different areas of the country. In NPMS children, homeless and institutionalised persons with psychiatric illness were not included, which may have led to a slight underestimation of prevalence rates. Because of the practical challenges in screening and gathering data, institutionalised, homeless, or imprisoned persons were not included in this survey. Variations in health systems between provinces, as well as sociocultural and economic conditions, may have had an impact on the outcomes at the national level. Because of the religious beliefs, family shame associated with suicidality, potential legal repercussions and overall stigma, it is possible that the rates of acknowledged suicidality and lifetime suicide attempts could be underreported.

### Conclusion

The results of the NPMS clearly shows that psychiatric disorders are a major public health issue with higher prevalence in middle-aged people, rural locations, lower income groups, and those with less education. The commonest psychiatric disorders are neurotic and mood disorders. It can be concluded that one out of every three adults in Pakistan suffers from a mental disorder. There is an urgent need to take steps to reduce this disease burden.

In response to the agenda 2030 of UNO for attaining SDG, the future mental health policy of Pakistan is moving towards the implementation of the bio-psychosocial model in which the psychological health is integrated into medical care. These policy initiatives set the trends for the next 5-Year Plan of Pakistan that is based on the principles of resources creation, community awareness, and establishment of feasible services that are affordable and accessible to all^68^. To facilitate the accomplishment of this policy, the NPMS provides sound evidence-based data about psychiatric disorders in Pakistan.

## Data Availability

All data produced in the present study are available upon reasonable request to the authors

## Acknowledgements

Team National Psychiatric Morbidity Survey (NPMS) is extremely thankful to the Pakistan Psychiatric Society and York University group for their financial support in carrying out the survey. We sincerely appreciate the advice provided by the National Expert Panel and National Technical Advisory Group members. Our sincere thanks to the Pakistan bureau of statistics and other government representatives for their valuable support during the various NPMS implementation phases. We would like to officially express our profound gratitude to every person and their families throughout Pakistan for their participation in the execution of the NPMS.

## Author contributions

All authors contributed to the design, planning and data collection for the study. The manuscript was drafted by Prof Raza ur Rahman. All authors critically reviewed and edited the draft manuscript for intellectual content and approved the final version after incorporating necessary changes. The corresponding author certifies that every listed author satisfies the requirements for authorship criteria and that no other qualified author has been left out. The group of partners for the National Psychiatric Morbidity Survey (NPMS) made data collecting possible in four Pakistani regions.

## Ethical approval

The National Bioethics Committee of Pakistan granted approval for this study vide letter # 4-87/NBC-268/171/370 dated November 11, 2017, and also by the Institutional Review Board of Dow University. Informed consent was obtained from each participant for this investigation. We attest that we have the proper permissions to use content protected by copyright.

